# The usefulness of SARS-CoV-2 test positive proportion as a surveillance tool

**DOI:** 10.1101/2020.07.06.20147843

**Authors:** Matt D.T. Hitchings, Natalie E. Dean, Bernardo Garcia-Carreras, Thomas J. Hladish, Angkana T. Huang, Bingyi Yang, Derek A.T. Cummings

## Abstract

Comparison of COVID-19 case numbers over time and between locations is complicated by limits to virologic testing confirm SARS-CoV-2 infection, leading to under-reporting of incidence, and by variations in testing capacity between locations and over time. The proportion of tested individuals who have tested positive (test positive proportion, TPP) can potentially be used to qualitatively assess the testing capacity of a location; a high TPP could provide evidence that too few people are tested, leading to more under-reporting.

In this study we propose a simple model for testing in a population experiencing an epidemic of COVID-19, and derive an expression for TPP in terms of well-defined parameters in the model, related to testing and presence of other pathogens causing COVID-19 like symptoms. We use simulations to show situations in which the TPP is higher or lower than we expect based on these parameters, and the effect of testing strategies on the TPP. In our simulations, we find in the absence of dramatic shifts of testing practices in time or between spatial locations, the TPP is positively correlated with the incidence of infection. As a corollary, the TPP can be used to distinguish between a decline in confirmed cases due to decline in incidence (in which case TPP should decline) and a decline in confirmed cases due to testing constraints (in which case TPP should remain constant). We show that the proportion of tested individuals who present COVID-19 like symptoms (test symptomatic proportion, TSP) encodes similar information to the TPP but has different relationships with the testing parameters, and can thus provide additional information regarding dynamic changes in TPP and incidence.

Finally, we compare data on confirmed cases and TPP from US states. We conjecture why states may have higher or lower TPP than average. We suggest that collection of symptom status and age/risk category of tested individuals can aid interpretation of changes in TPP and increase the utility of TPP in assessing the state of the pandemic in different locations and times.

**Summary:**

- Key question: when can we use the proportion of tests that are positive (test positive proportion, TPP) as an indicator of the burden of infection in a state?
- If testing strategies are broadly similar between locations and over time, the TPP is positively correlated with incidence rates.
- However, changes in testing practices over time and between locations can affect the TPP independently of the number of cases.
- More testing of asymptomatic individuals, e.g. through population-level testing, lowers the TPP.
- We can identify locations that have a lower or higher TPP than expected, given how many cases they are reporting.
- Efficient transmission increases detected cases exponentially, resulting in large changes in confirmed cases compared to factors that change linearly.
- Data that could aid interpretability of the TPP include: age of individuals who test positive and negative, and other data on testing performed in high-prevalence settings; and symptom status of tested individuals.

## Introduction

The number of confirmed SARS-CoV-2 infections is determined by the true infection rate and the number and type of people who are tested for presence of the virus. Monitoring the pandemic in a location (or comparing locations) by the number of reported cases is confounded by the amount and type of testing being done; the number of cases is inherently limited by the number of tests performed. The test positive proportion (TPP) has been used as an additional indicator of case; a high TPP coupled with a significant number of cases is seen as an indication that the reported cases represent the “tip of the iceberg” and that testing capacity should be increased to get a better understanding of transmission. Where capacity is limited, tests are given preferentially to those more likely to be positive, such as hospitalized patients, meaning that mildly symptomatic infections (i.e. the majority of infections) are likely to be undetected. On the other hand, a low TPP is viewed as an indication of a potentially effective surveillance and containment strategy^1^, and implies that increased testing would not reveal a significant number of undetected infections. The WHO and US CDC have made TPP part of their guidelines for easing lockdown restrictions, indicating that it can be used to assess readiness for releasing restrictions and recommending that communities should be below 5% for 14 days before they consider relaxing social distancing measures^2,3^.

While these interpretations are broadly plausible, many aspects of testing strategies, including the rate of testing of symptomatic and asymptomatic individuals, the number of tests available, and the performance of the test, could change TPP independently of the true infection rate. We aim to explore the relationship between TPP and testing parameters, and suggest additional metrics and data to aid interpretation of the TPP.

## Method

### Model for disease and testing

We model transmission of SARS-CoV-2 in a population using an SEIR (Susceptible, Exposed, Infectious, Recovered) model (see Supplementary Appendix for model equations and further details). The state variables represent the number of individuals in each compartment, with *S* + *E* + *I* + *R* = *N*. All individuals in the *I* compartment are infectious and have COVID-19 like illness (CLI) until they recover. A proportion *p*_*I*_ of those in the *S* and *E* compartments have CLI from other causes (henceforth “non-SARS-CoV-2 CLI”).

We model the application of tests using compartments for available test kits and completed tests applied to SARS-CoV-2 positive/SARS-CoV-2 negative and symptomatic/asymptomatic individuals. We assume that an individual’s rate of testing differs by symptom status. Recovered individuals are assumed not to seek testing. The test has sensitivity *p*_*S*_. Exposed and infectious individuals are quarantined upon testing (efficacy of quarantine is assumed to be perfect, represented by moving those individuals to the R compartment).

A schematic for the natural history and testing model is shown in Figure 1 (see Supplementary Appendix for a table of parameters). The demand for tests is equal to the number of individuals in each group multiplied by the rate of test-seeking. Let *d*_*A*_and *d*_*S*_ be the daily rate of testing among asymptomatic and symptomatic individuals. For example, every day a proportion d_S_ of symptomatic individuals will receive a test. Total demand *D* = (*p*_*I*_*d*_*S*_ + (1 − *p*_*I*_)*d*_*A*_)(*E* + *S*) + *d*_*S*_*I* (i.e. the total number of tests sought by the entire population each day). The demand from any specific group is denoted using subscripts, e.g. *D*_*ES*_ for exposed individuals with CLI. We assume that anyone with CLI seeks testing at the symptomatic rate d_S_.

**Figure 1:**
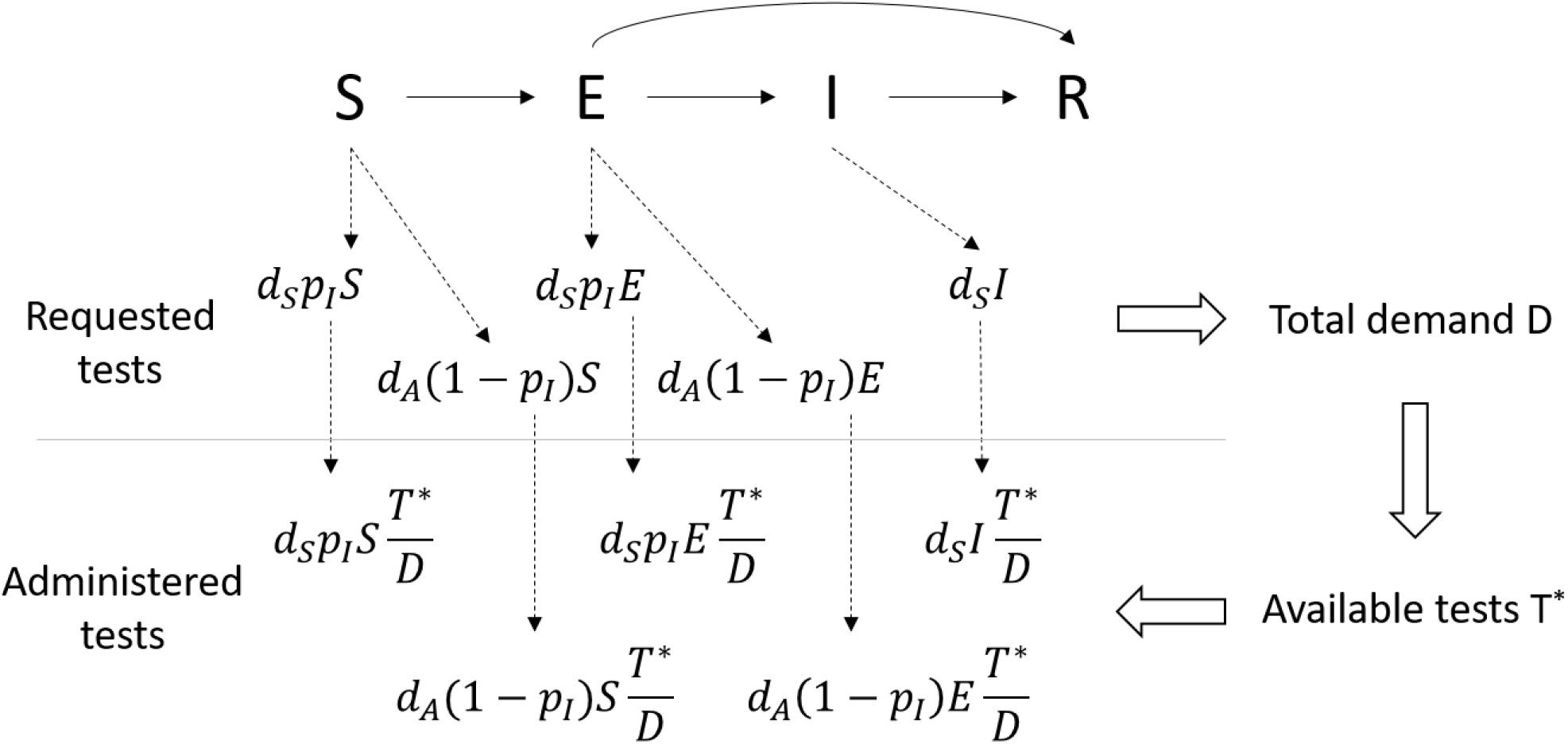
Schematic for disease natural history (SEIR) and testing. Requested tests from each compartment are determined by the proportion of individuals with COVD-19 like symptoms in each compartment, and by the rate of testing among symptomatic and asymptomatic individuals. Tests administered each day are limited by available tests, and assigned proportionally to each compartment according to demand.

The number of tests performed per day is limited by a daily maximum number of tests that can be performed *T*_*max*_ (i.e. laboratory capacity), and by the number of available test kits *T* (i.e. test stockpile). We choose a simple functional form for the number of tests performed per day: *T*^∗^ = *min*(*T, D, T*_*max*_).

If demand for testing exceeds the number of available test kits, the rate of testing for each compartment is normalized by the demand (e.g. rate of testing among symptomatic, exposed individuals is *D*_*ES*_*T*^∗^/*D*). Thus, test kits are allocated to each type of individual proportional to demand.

### Relationship between incidence and TPP

The above model gives rise to an equation for TPP as a function of infectious prevalence and the model parameters. We assume that early in the epidemic, there are few individuals in the R compartment (*R* ≈ 0). The TPP is the number of positive tests over the total number of tests carried out on a given day, or

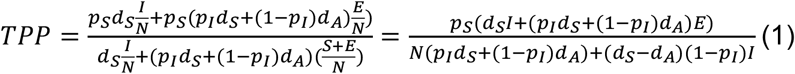

as *S* + *E* = *N* − *I* when *R* is very small. Rearranging gives

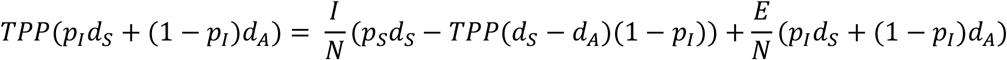

All components of the above equation are positive if *d*_*A*_ ≤ *d*_*S*_. An upper bound for infectious prevalence is thus

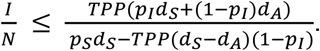

If the *E* class contributes little to detected cases compared to the *I* class (i.e. *p*_*I*_*d*_*S*_ + (1 − *p*_*I*_)*d*_*A*_<< *p*_*S*_*d*_*S*_ − *TPP*(*d*_*S*_ − *d*_*A*_)(1 − *p*_*I*_)), the above inequality becomes an approximate equality.

Given the infectious prevalence *I*/*N* at a point in time and assuming that the majority of detected cases are from the infectious compartment, the number of positive tests per capita (henceforth “confirmed incidence”) is estimated by 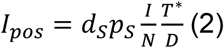. We can write TPP as a function of the confirmed incidence rate,

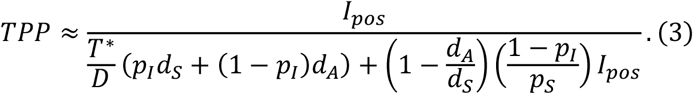

The model allows us to examine the effect on TPP of changes in the parameters that determine testing *T*^∗^. If the relevant parameters were known we could use equation (2) to “estimate” the confirmed incidence from the TPP; we refer to this as the “TPP-estimated” incidence.

### Additional metrics to aid interpretation of TPP

If the number of symptomatic individuals among those seeking testing is known, the test symptomatic proportion (TSP) provides a similar relationship between confirmed incidence and TSP, assuming *R* ≈ 0:

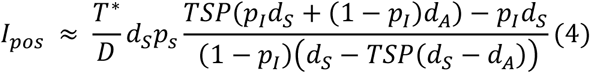

If the population contains a high-risk group, with increased susceptibility to infection and thus higher infectious prevalence, the overall TPP will depend on the proportion of tested individuals in the high-risk group. Therefore, we assume that the groups are tested at different rates and stratify the TPP and TSP by risk group.

### Assessment of TPP-incidence and TSP-incidence relationships using simulations

As the above equations involve approximations and changes in infectious prevalence over time cannot be derived analytically, we assess the accuracy of the equations using simulation. We simulate the SEIR model described above using an adaptive tau-leaping method (R package adaptivetau). From the simulations we calculate the weekly confirmed incidence, TPP, TSP, and supply/demand ratio (stratified by susceptibility group where relevant).

We assess the relationship between TPP, TSP and confirmed incidence, compared the relationship in Equations (3) and (4), varying testing parameters d_A_, d_S_, and p_I_. We then explore the impact of dynamic changes of model parameters in time by inducing linear changes in parameters in time. Finally, to understand the effect of having a high-susceptibility group (e.g. essential workers) in the population, with possibly higher rates of testing, we vary the proportion of the population that is high-risk, the relative susceptibility, and relative rates of testing between the high- and low-risk groups.

### Work with empirical data

In addition to our simulations, we use data from the COVID Tracking Project^4^ to show the relationship between TPP and confirmed cases across states, and within states across time, with population data for states from the US Census Bureau^5^. We use the derived equations to plot the expected relationship between TPP and confirmed cases if all states had the same testing parameters, by fitting equation (2) to the observed confirmed cases using ordinary least squares (OLS) regression. At a point in time, we can identify states that have higher TPP than average among states with similar incidence, and vice versa, and provide intuition for why those states might have different TPP than average. Similarly, we fit equation (2) to observed confirmed cases for a time series within a single state to identify periods of time in which the trend in TPP is not expected given the trend in confirmed cases. In addition to national data, we use data from the Oregon Health Authority^6^, which reports the proportion of COVID-19 cases with various symptoms to illustrate trends in symptoms over time.

## Results

### Relationship between TPP and confirmed incidence

The univariate relationship between TPP and the model parameters is shown in Figure 2. We varied each parameter in turn, with the other parameters fixed at default values (see Supplementary Appendix). TPP increases as the rate of confirmed infections increases (A), as the prevalence of infectious individuals is higher and consequently there is a greater demand for tests from symptomatic, SARS-CoV-2 positive individuals. Testing strategy can affect the TPP and TSP. Counter-intuitively, if symptomatic individuals are tested at a higher rate the TPP will decrease (B) if the rate of confirmed infections is held constant. In this case, higher rate of testing symptomatic individuals means that confirmed infections represent a larger proportion of all infectious individuals, and infectious prevalence decreases with higher rate of testing symptomatic individual. Finally, lower infectious prevalence translates to lower TPP. If more asymptomatic individuals are tested (e.g. through expanding eligibility for testing), the TPP will decrease (C). If there is a shortfall of testing supply relative to demand (D), TPP will increase because the confirmed cases represent a smaller proportion of infectious individuals, and more overall demand from infectious individuals leads to higher TPP. Finally, factors independent of policy decisions can affect the TPP. The test sensitivity has a negligible effect on the TPP (E) when the confirmed incidence is held constant. On the other hand, if prevalence of non-SARS-CoV-2 CLI is higher (e.g. during an influenza outbreak), the TPP will decrease as more SARS-CoV-2 negative individuals will seek testing (F).

**Figure 2:**
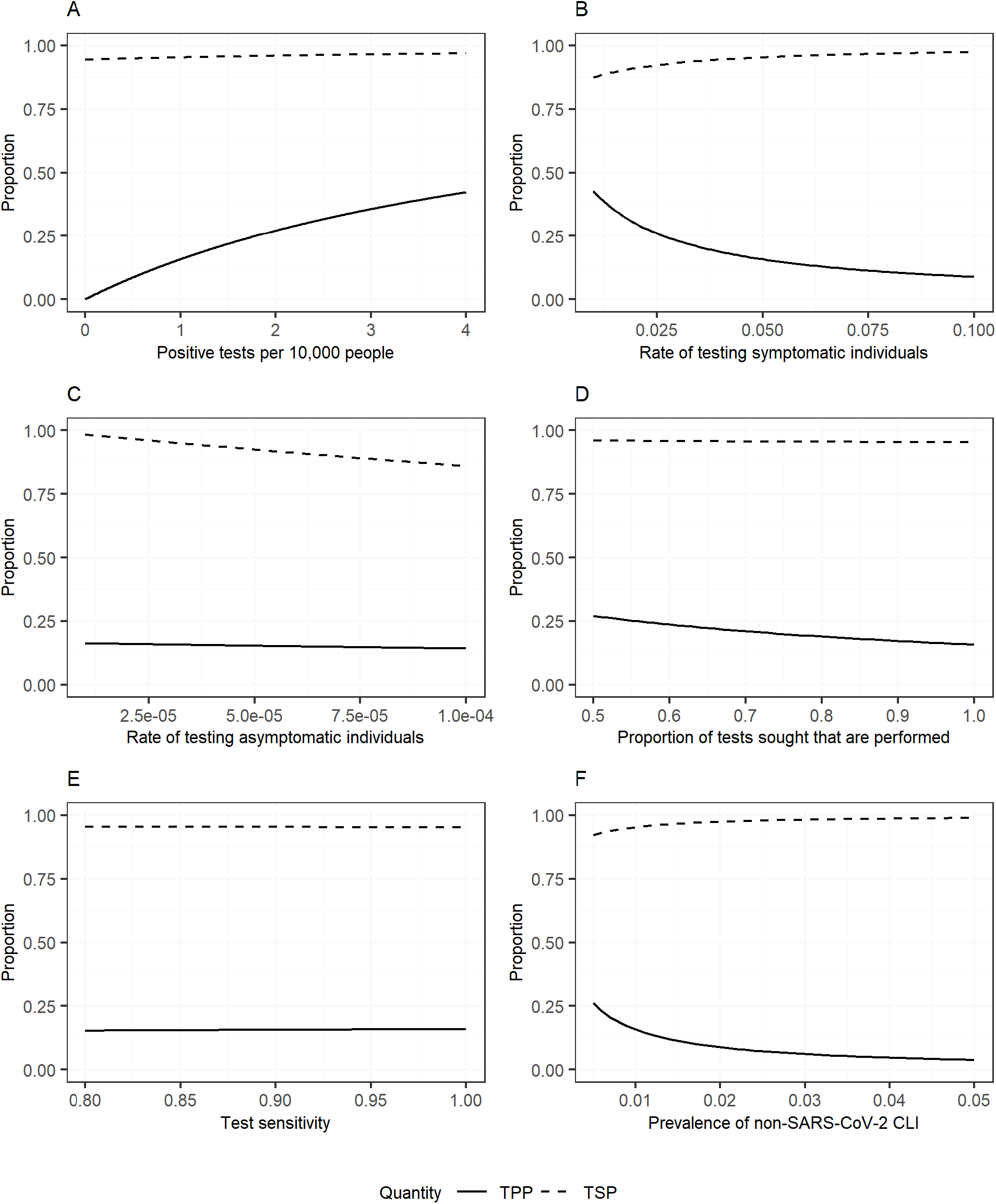
Relationship between TPP (solid line) and TSP (dotted line) and rate of incident confirmed infections of SARS-CoV-2 (A), rate of testing among symptomatic individuals (B), rate of testing among asymptomatic individuals (C), shortfall of test supply relative to demand (D), test sensitivity (E), and prevalence of non-SARS-CoV-2 CLI (F).

Figure 2 also shows the relationship between TSP and the model parameters. TSP increases modestly as infectious prevalence increases (A), and decreases as the testing rate among asymptomatic individuals increases (B). In contrast to the TPP, the TSP increases with higher testing rate of symptomatic individuals (C) and higher prevalence of non-SARS-CoV-2 CLI (D), as both of these parameters lead to increased testing demand from symptomatic individuals. Testing supply shortfall (E) and test sensitivity (F) have negligible effects on the TSP.

equation (2) provides intuition for how TPP and detected cases will change under different scenarios. During the exponential growth phase of an epidemic, TPP will rise as infectious prevalence rises rapidly (Figure 2A). Similarly, if the rate of new infections is declining and testing strategies remain constant, TPP will decrease over time. This observation provides a simple way to understand whether falling case numbers is due to a true decline in incidence or a shortage of test kits. If the infectious prevalence declines, the rate of confirmed cases and the TPP will both decline (Figure 2A), whereas if the supply of test kits is falling relative to demand but the infectious prevalence remains constant, the rate of confirmed cases will decrease (equation (2)) but the TPP will remain constant (Equation (1); if infectious prevalence is constant then TPP is independent of supply of test kits). Similarly, rising TPP concurrent with rising confirmed incidence is an indication that infectious prevalence is truly rising. If a rise in confirmed cases were due to increases in testing capacity alone, we would not expect TPP to increase.

### Relationship between TPP and TSP

Figure 1 demonstrates how the TSP could provide further information to interpret changes in confirmed cases and TPP. The relationship between TSP and testing parameters is in some cases the inverse of the relationship between TPP and the parameters. For example, a decrease in TPP coupled with a rise in TSP over time provides evidence for an increase in prevalence of non-SARS-CoV-2 CLI, or in the rate of testing among symptomatic individuals, over a change in testing rate among asymptomatic individuals. External data on changes in ILI over time can further narrow down the cause of dynamic changes in TPP and TSP.

### Simulation results

We assessed the accuracy of the TPP and TSP formulas using data generated from an SEIR model. We fixed the testing parameters for all simulations and sampled pre-lockdown R_0_ uniformly from 3 to 5, time of lockdown from 21 to 35 days, and post-lockdown R_0_ from 0.8 to 1 in the absence of testing. Figure 3 shows that if the supply of test kits is sufficient to cover the demand from symptomatic and asymptomatic individuals, the relationship between TPP and confirmed incidence is as in equation (2) (blue points), but that if there is limited supply of test kits the proportion of cases detected decreases while the TPP remains the same. Therefore, the TPP observed in the simulations are greater than predicted (red points). We observe a similar pattern for TSP.

**Figure 3:**
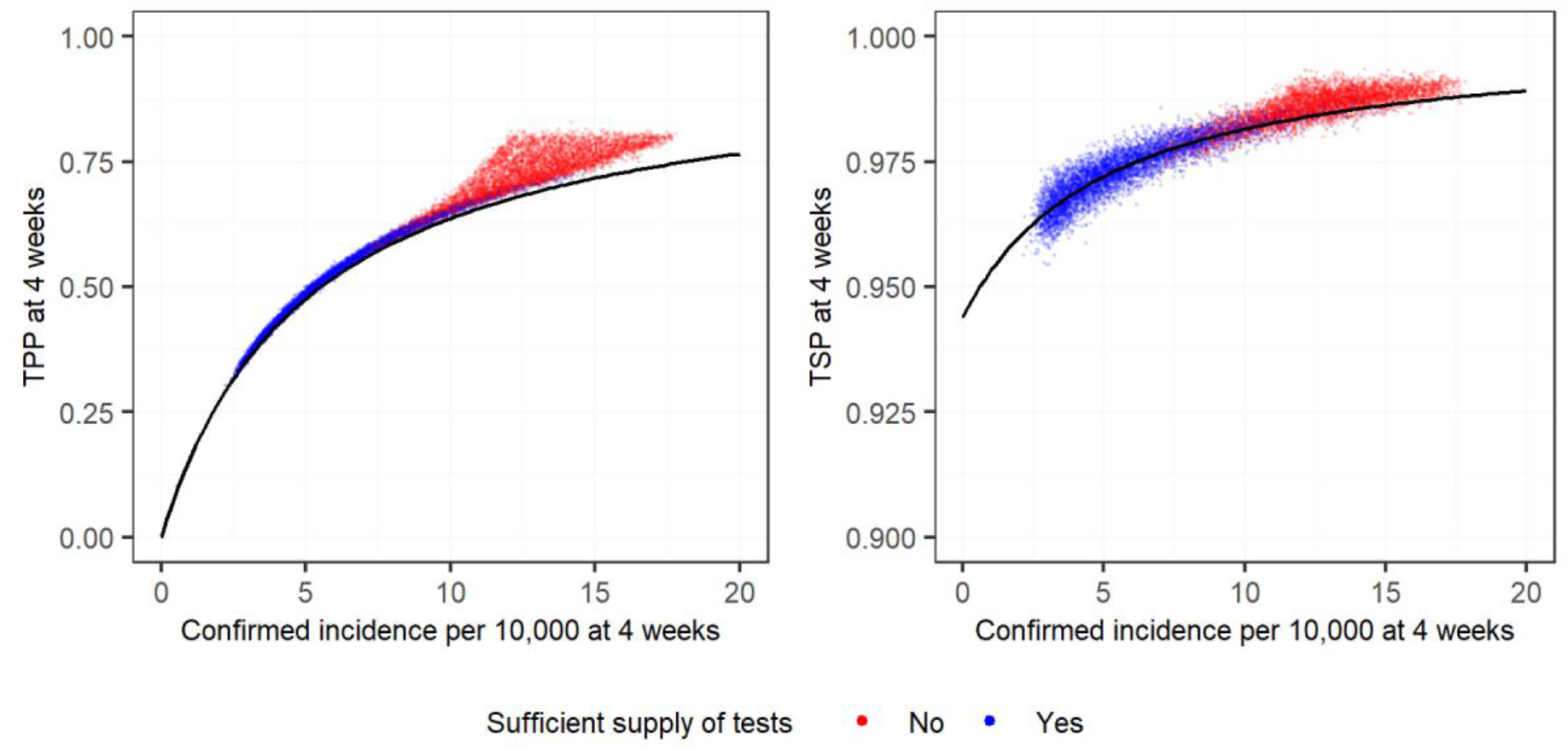
Confirmed incidence per 10,000 against TPP (left) and TSP (right) at 4 week, from 10,000 SEIR simulations in which initial and lockdown R_0_ values are randomly sampled from specified ranges (see text). The color of the points represents whether supply of testing was sufficient to cover demand at week 4, which is a function both of the availability of test kits and R_0_. The solid line in each panel denotes the relationship in Equations (3) and (4).

Across simulations, TPP was strongly correlated with the confirmed incidence rate (average Pearson correlation = 0.94 across all simulations in Figure 3). Correlation remained high in the presence of linear changes in testing parameters over time within a location (see Supplementary Appendix for more details). Simply put, the correlation between detected cases and TPP during the growth phase is broken only if the number of tests performed increases more quickly than the number of positive tests. (See Supplementary Appendix for results including a high-susceptibility group with differential testing).

### Comparison of TPP and confirmed incidence across US states

Figure 4 shows the relationship between confirmed incidence per 10,000 and TPP by state, at four different times relative to the start of the epidemic in each state. The parameter values that minimize the sum of squares are plotted as a regression line. If all states had the same testing parameters and sufficient supply of test kits, we would expect them to lie on the line as in Figure 3 (blue dots).

**Figure 4:**
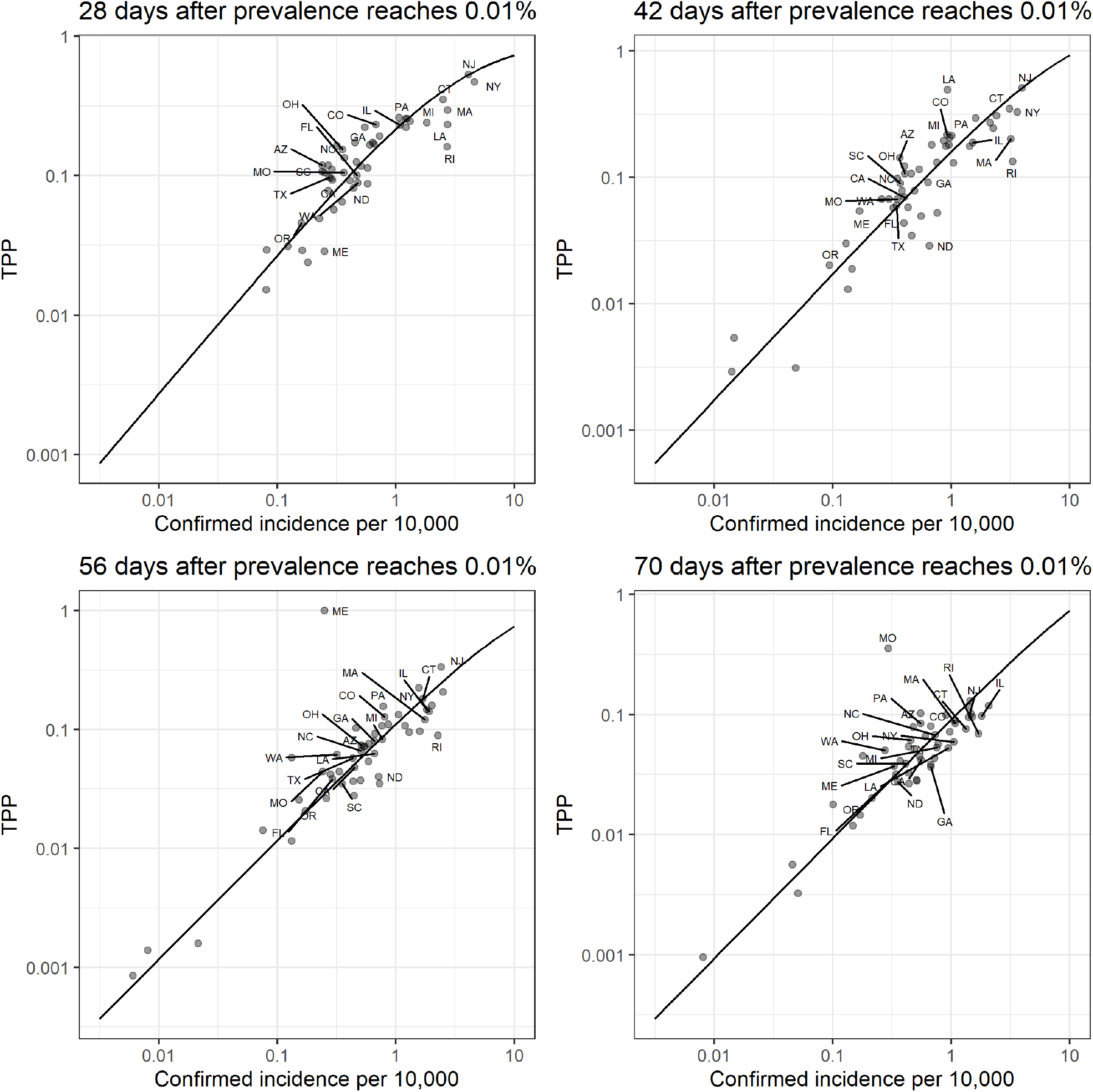
Weekly average of observed confirmed incidence (x-axis) vs TPP (y-axis) on a log-log scale. The trend line is fit using equation (2) for each time point separately.

States that fall below the line have a lower TPP than would be expected for how many cases they have observed. Figure 2 shows that there could be several reasons for this: increased prevalence of non-SARS-CoV-2 CLI; increased testing of asymptomatic individuals; or increased testing of symptomatic individuals and correspondingly lower infectious prevalence. States that fall above the line have a higher TPP than would be expected for how many cases they have observed. The reasons for this are the inverse of those above. In addition, as in Figure 3, there could be a shortfall of supply of test kits relative to demand, leading to a lower rate of case detection than the average state. We observe a linear relationship between confirmed incidence and TPP, except in the first panel where there is some evidence of saturation of TPP at high incidence rates experienced early in the epidemic. Most states fall close to the average, while three states (Massachusetts, Rhode Island, and North Dakota) appear to have consistently had lower-than-average TPP. On the other hand, Colorado appears to have consistently had higher-than-average TPP. Outliers (Louisiana, Maine, and Missouri in panels 2, 3, and 4) are explained by changes in reporting of test data (for example, reporting serology and PCR tests separately that had previously been reported together)^4^.

Figure 5 shows time series for individual states: confirmed 7-day rolling average confirmed incidence (solid line) and “TPP-estimated” 7-day average confirmed incidence (dotted line), with parameters for each state estimated using OLS. Figure 5 demonstrates that often the TPP (and thus TPP-estimated incidence rate) correlates with observed incidence. Periods of time when this is not the case are indicative of dynamic changes in testing strategy leading to tests being targeted more towards positive or negative individuals, a change in supply of test kits relative to demand, or a change in reporting of tests in the data that may not represent changes in the number of tests being administered.

**Figure 5:**
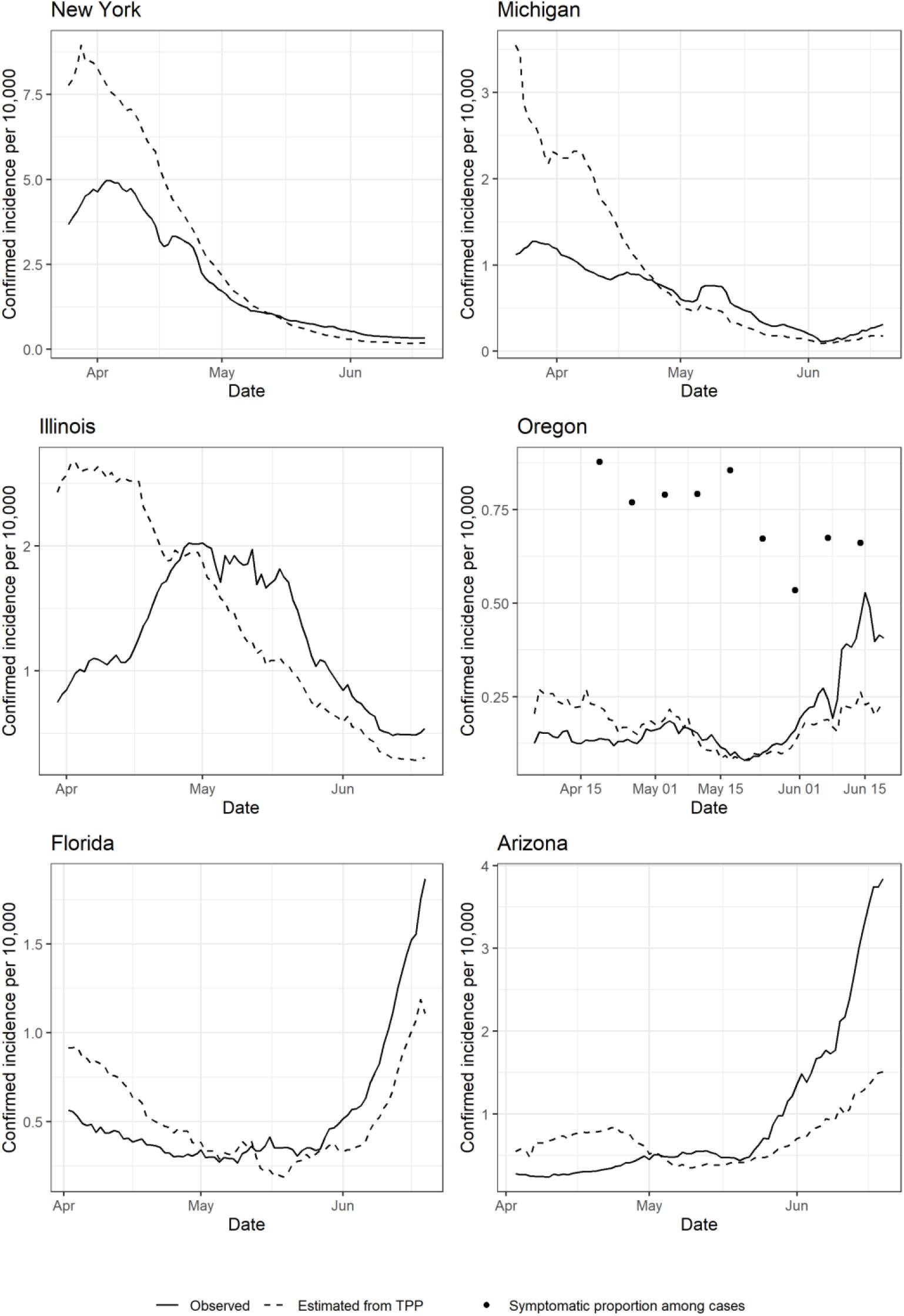
7-day rolling average confirmed incidence (solid line) and “TPP-estimated” 7-day average confirmed incidence (dotted line). Parameters for estimated incidence are fitted to each time series separately. In the fourth panel, points represent the proportion of confirmed cases that were symptomatic, plotted on the same axis.

For example, in early April the TPP in New York and Michigan was higher than in May and June, adjusting for the observed incidence rate, due to tests being given preferentially to sicker or higher-risk individuals. In Illinois, the rise in confirmed cases in April was accompanied by a decrease in the TPP, suggesting an expansion of testing capacity and a systematic targeting of healthier individuals. In Oregon, confirmed incidence has risen in June while TPP has risen less dramatically. In addition, the proportion of confirmed cases reporting symptoms decreased from May to June. Although not the TSP as we have defined it, this metric gives an indication of how much testing is being conducted among asymptomatic individuals. In Florida and Arizona (bottom row), the rise in cases in June is accompanied by a rise in TPP, and thus TPP-estimated incidence rate, as we would expect if infectious prevalence were increasing.

## Discussion

We present a simple model for testing for SARS-CoV-2 within an SEIR framework to derive an expression for TPP as a function of well-defined parameters. We use this expression to understand how TPP changes with the confirmed incidence as well as other parameters related to testing and the presence of other pathogens in the population. In particular, our work can be used to build hypotheses for why a location or point in time has a higher/lower TPP than expected.

When comparing TPP between locations, it is important to compare the rate of incident confirmed cases at the same time. Within the US, earlier in the epidemic New York and New Jersey were pointed to as examples of states that had very high TPPs compared to the country average, but this analysis shows that they are in line with the average after adjusting for the observed incidence.

A key question in locations in which confirmed incidence is increasing is whether this growth is due to a true increase in infectious prevalence. An alternative explanation of increased testing capacity coupled with more tests being applied to populations at higher risk of infection might explain both the increasing number of cases and the increasing TPP. Increased testing in groups with higher prevalence of infection will increase the TPP, but sustained rises in TPP concurrent with rises in case numbers are easily explained by rising infectious prevalence in the population. Publicly available data on testing in different high-risk populations is critical in teasing apart these hypotheses. To our knowledge, the CDC website is the only source of such information stratified by age in the US^7^, and this information is only available at a single time point. The patterns shown there are striking and merit further investigation: namely that decreases in TPP in April and May were driven primarily by decreases in TPP among those >50 years old.

Other authors have attempted to infer the population prevalence in the US using case data and TPP^8^. In this analysis we use a mechanistic model to explain the apparent linear relationship observed between confirmed incidence rate and TPP across locations. We expect the relationship to be non-linear, tending asymptotically to TPP=1 as the prevalence of infectious individuals increases. However, this occurs at higher infectious prevalence than is observed in the data we have used, meaning that the model is not identifiable as we have five parameters to fit a single linear gradient.

The assumption that allocation of test kits is proportional to demand implies that selection bias in the sample of individuals tested is independent of the number of test kits available. It may be that in the cases of extreme restriction in testing availability, more priority is given to sicker individuals seeking testing. Therefore, deviations from the expected TPP when testing availability is limited would be more extreme than observed in our simulations.

The disease model presented here is a simplification of the true natural history, ignoring individuals who are asymptomatic but infectious during the course of infection, and pre-symptomatic infectiousness. Accounting for these types of individuals would increase the contribution of asymptomatic individuals to the TPP, likely making the “TPP-estimated” incidence less accurate. However, the observation that TPP should theoretically increase monotonically with infectious prevalence remains. In addition, we have simplified the distinction between “high” and “low” susceptibility groups. In reality, the elderly and those with comorbidities form a high-risk group that have increased probability of disease given infection^9^. In addition, other groups might be at higher risk of exposure and thus infection (e.g. essential workers) but not at higher risk of disease. Increased symptomatic disease among those with more risk of exposure (and thus likely higher prevalence of infection) will lead to increased testing in this group. This will exacerbate the bias in the overall TPP caused by the existence of the different risk groups.

In conclusion, we have provided intuition for how TPP changes in response to the prevalence of infectious individuals in the population as well as with various testing parameters. Unless infectious prevalence is extremely high, we expect a linear relationship between detected incidence rate and TPP across locations, and deviations from this relationship can be interpreted using the model equations. In general, if a location has higher TPP than expected, this means testing is targeted more towards sicker individuals, or there is a shortage of test kits relative to demand. As case counts begin to increase in several US states, increasing TPP should be interpreted as a warning sign that transmission is increasing. TPP stratified by age or other indicator of risk group will aid interpretation of changing case numbers.

## Data Availability

R code for simulations provided as Supplementary Material. All data used in study is publicly available, with links provided in manuscript.

